# The Emerging Applications of Synthetic Data in Neurosurgery Research and Practice: A Systematic Review

**DOI:** 10.1101/2024.03.04.24303724

**Authors:** Andrew Bouras

## Abstract

**Background:** The integration of data-driven technologies into neurosurgery, particularly through the advent of synthetic data, marks a significant evolution in the field. This qualitative systematic review explores the impact of synthetic data on neurosurgical practices, including preoperative planning, intraoperative navigation, postoperative care, training, and research. The goal is to provide a comprehensive assessment of the current applications, benefits, challenges, and future directions of synthetic data in neurosurgery.

**Methods:** A thorough literature review was conducted, focusing on peer-reviewed articles and conference proceedings that detail the use of synthetic data in neurosurgery. The review prioritized studies offering qualitative evaluations, case studies, technological developments, and expert perspectives on synthetic data’s integration into neurosurgical practices. Inclusion criteria were established to select studies that explicitly discuss the generation, utilization, and impact of synthetic data in the discipline.

**Results:** The analysis reveals that synthetic data contributes significantly to neurosurgery, enhancing surgical planning precision, training simulation accuracy, and enabling personalized care. Identified benefits include addressing the scarcity of clinical data, maintaining patient privacy, and facilitating machine learning model development. Challenges such as ensuring data realism and variability, along with the integration of synthetic data into clinical workflows, were also identified. The review further highlights synthetic data’s role in supporting collaborative research, navigating data sharing obstacles, and fostering innovation in neurosurgical methods and patient outcomes.

**Conclusions:** Synthetic data presents a transformative opportunity for neurosurgery, addressing historical challenges and fostering advancements. Despite existing hurdles, its application across neurosurgical domains indicates a shift towards more personalized, precise, and effective patient care. Overcoming these challenges requires ongoing interdisciplinary cooperation and technological progress. The review emphasizes the necessity for standardized methodologies, ethical considerations, and a proactive stance to leverage synthetic data’s potential fully in neurosurgical advancements.

## 1 Introduction

### 1.1 Background on increasing role of data in advancing neurosurgery

The journey of neurosurgical care begins well before the operating room, where meticulous planning is paramount. In recent years, the utilization of patient-specific anatomical models and virtual reality simulations has transformed preoperative planning. These tools, enriched by predictive analytics, allow surgeons to forecast surgical outcomes with greater accuracy, enabling tailored interventions that consider the unique physiological and anatomical nuances of each patient. This personalized approach is critical for minimizing risk and optimizing surgical strategies, setting the stage for successful outcomes.

The complexity of neurosurgical procedures demands precision and adaptability, challenges that are met with the advent of intraoperative navigation systems. Integrating multimodal imaging and biomechanical modeling with augmented reality, these systems offer real-time, dynamic guidance to surgeons. By compensating for the inevitable brain shift and other anatomical changes during surgery, they ensure that interventions remain precise and aligned with preoperative plans, thereby enhancing the safety and effectiveness of the surgery.

The role of data extends into the postoperative phase, where analytics play a crucial role in shaping patient recovery. By analyzing trends from both individual patient data and broader population-level evidence, healthcare providers can make informed decisions regarding follow-up care, rehabilitation, and potential interventions. This data-driven approach ensures that postoperative care is as personalized and effective as the surgery itself.

Despite these advancements, the neurosurgical field continues to face significant challenges related to the availability, quality, and diversity of clinical data. These limitations often hinder the development and application of innovative technologies, prompting a search for alternative solutions. Synthetic data generation and simulation have emerged as promising avenues to address these gaps, offering a means to create detailed, versatile datasets that can support the continuous evolution of neurosurgical techniques and technologies.

Looking forward, the integration of synthetic data into neurosurgical practice holds the potential to revolutionize the field. By providing a rich substrate for training, planning, and procedural assistance, synthetic data can help overcome existing barriers, paving the way for further innovations that enhance patient outcomes. As the neurosurgical community embraces these new tools, the focus remains on harnessing the power of data to refine surgical precision, improve patient care, and navigate the complexities of the human brain with unprecedented insight.

In sum, the trajectory of neurosurgery is increasingly data-driven, marked by significant technological advancements that expand the possibilities of what can be achieved in patient care. As the field continues to evolve, the thoughtful application of both real and synthetic data will be crucial in unlocking the next frontier of neurosurgical innovation.

### 1.2 Challenges around limitations of real world clinical data

One of the most formidable obstacles is the presence of data silos, where valuable clinical information is confined within the boundaries of individual institutions. Privacy regulations, consent protocols, and local governance policies, although crucial for protecting patient information, often complicate the sharing and aggregation of data across healthcare systems. This fragmentation hampers the ability to perform comprehensive analyses that require large, diverse datasets, particularly for developing and training advanced machine learning algorithms tailored to neurosurgery.

Within healthcare institutions, the challenge of interoperability between different data systems further exacerbates the difficulty of conducting holistic analyses. These technological and systemic limitations create information gaps that can leave significant insights untapped, diminishing the potential for data-driven improvements in neurosurgical care.

The detailed annotation of complex surgical imaging datasets is essential for training deep learning models to recognize and interpret intricate pathological features. However, the scarcity of professionals with the requisite expertise for such meticulous work introduces a bottleneck in the data preparation process, making large-scale dataset labeling both time-consuming and costly.

Variability in data collection protocols and inherent sampling biases can lead to datasets that poorly represent rare but critical clinical phenomena. This issue is particularly problematic when dealing with niche subpopulations or complex cases where specific data points are crucial for accurate analysis and decision-making.

Adhering to strict de-identification standards to ensure patient privacy further complicates the sharing of data for retrospective studies and research, limiting the ability to leverage existing clinical data for broader analyses that could inform future neurosurgical practices.

These multifaceted challenges highlight the growing importance of exploring alternative approaches, such as synthetic data simulation, to circumvent the limitations of real-world clinical data. By generating high-quality, representative datasets through synthetic means, the neurosurgical field can gain access to a broader array of data for innovation, training, and research. Synthetic data not only offers a pathway to overcoming data scarcity and privacy concerns but also presents an opportunity to standardize data for more consistent and comprehensive analyses.

### 1.3 Introduction to synthetic data and its applications

At its core, synthetic data is crafted through algorithms that replicate the statistical properties of real-world data, achieving a high degree of similarity to actual clinical datasets. This replication process is designed to sidestep the myriad challenges related to data access, privacy concerns, and the ethical use of sensitive patient information, which traditionally hinder the aggregation of diverse data sets necessary for comprehensive analysis and innovation in healthcare.

The creation of synthetic data addresses critical issues by enabling the generation of extensive datasets that mirror the complexity of real patient data. This breakthrough facilitates research and development activities that were previously constrained by the scarcity of accessible data, especially concerning rare conditions or unique patient populations. By automating the generation process, synthetic data allows for the creation of large-scale datasets that encompass a broad spectrum of data types, from common occurrences to rare phenomena, thus significantly enhancing the scope of medical research and application.

Beyond increasing data availability, synthetic data generation techniques incorporate mechanisms to minimize biases and inaccuracies, thereby ensuring the data’s reliability and relevance for medical applications. These generative models are meticulously designed to maintain the integrity and authenticity of clinical information while embedding layers of anonymity to protect patient privacy. This balance enables the safe and ethical distribution and sharing of data across research institutions and healthcare providers, facilitating collaborative efforts in medical innovation.

The advent of synthetic data has opened up new avenues for advancement across various neurosurgery subdomains. Its applications range from refining predictive analytics and facilitating personalized surgical planning to enhancing the accuracy of intraoperative navigation systems. Furthermore, synthetic data plays a crucial role in the development of simulation-based training programs, providing a risk-free environment for surgeons to hone their skills and adapt to new technologies. The use of synthetic datasets also supports the generation of evidence for data-driven decision-making processes, thereby improving the quality of patient care and outcomes.

As the technology behind synthetic data continues to evolve, its integration into neurosurgical practices promises to bridge the gap between current limitations and the potential for groundbreaking innovations. The ability to generate detailed, accurate, and privacy-compliant datasets positions synthetic data as a cornerstone technology that will shape the future of neurosurgery. With ongoing advancements in deep learning and computational modeling, synthetic data is transitioning from a concept of simple data replication to a comprehensive tool capable of producing highly sophisticated digital twins of clinical realities.

## 2 Methods

### 2.1 Search strategy

A comprehensive search of scientific databases, including PubMed, IEEE Xplore, Google Scholar, and ScienceDirect, was performed to gather literature relevant to the use of synthetic data in neurosurgery. Search terms such as “synthetic data,” “neurosurgery,” “predictive analytics,” “surgical planning,” “machine learning,” and “data simulation” were employed to capture pertinent studies. The literature review spanned from January 2000 to February 2024, focusing on recent advancements. The initial selection process involved screening titles and abstracts for relevance to synthetic data applications and machine learning (ML) techniques in neurosurgical settings. This was followed by full-text reviews to evaluate clinical efficacy, safety, and the role of ML in enhancing neurosurgical procedures with synthetic data. Priority was given to high-quality primary research published in peer-reviewed journals.

### 2.2 Eligibility criteria

Studies were excluded if they were conference papers, reviews, lacked a primary research focus, or addressed topics not directly related to synthetic data in neurosurgery. Data extraction involved recording the objectives, methods, key findings, and conclusions of each study, enabling a qualitative synthesis of trends and themes related to the integration of synthetic data and ML in neurosurgery. The critical appraisal of each study’s limitations and potential biases provided a balanced and comprehensive perspective, informing future research directions in the field.

### 2.3 Study selection and data extraction

The selection process rigorously assessed studies for their contribution to integrating synthetic data with neurosurgical practices. Analysis centered on objectives, methods, outcomes, and implications, aiming to elucidate the transformative role of synthetic data in neurosurgery. This approach enabled the identification of key trends, patterns, and themes, particularly highlighting how synthetic data and machine learning technologies enhance surgical planning, execution, training, and the personalization of patient care.

### 2.4 Study Quality Assessment

Each study underwent a critical evaluation for methodological rigor, potential biases, and limitations, ensuring the review’s integrity and reliability. This assessment aimed to present an unbiased overview of the current research landscape, highlighting strengths and areas for improvement in the literature on synthetic data and ML in neurosurgery.

### 2.5 Data analyses

A qualitative synthesis was chosen as the method for analyzing the collected data, providing a comprehensive understanding of the complex roles and impacts of synthetic data and ML in neurosurgery. Special attention was paid to the ways in which these technologies have influenced surgical planning, execution, training, and overall patient care outcomes.

## 3 Results

### 3.1 Overview of synthetic data for machine learning in neurosurgery

The pioneering work of Lindner et al. (2019) and Pantovic et al. (2022) exemplifies the transformative potential of synthetic data in neurosurgery. Lindner and colleagues have demonstrated the successful training of a glioblastoma detection model using synthetic MRI data generated by generative adversarial networks (GANs). Remarkably, this model achieves accuracy comparable to those trained on real datasets, despite the synthetic nature of its training images. This achievement not only underscores the viability of synthetic data to substitute real images in training deep learning models but also highlights the potential for significant reductions in data acquisition and annotation costs.

Similarly, the research conducted by Pantovic et al. introduces an innovative approach to improving the robustness of CT scan analyses. By integrating synthetically generated data that simulates metal artifacts into the training process, their model demonstrates enhanced capability in accurately identifying crucial surgical landmarks, such as intracranial electrodes. This methodological advancement signifies a crucial step towards overcoming the limitations posed by artifact interference in clinical imaging, paving the way for more reliable surgical planning and outcomes.

These studies collectively signal a growing recognition of synthetic data as a powerful tool in the field of medical imaging and neurosurgery. By enabling the training of deep learning models on diverse and complex datasets without the traditional barriers of data scarcity and high costs, synthetic data opens new avenues for the development of advanced diagnostic tools. This progress is particularly relevant for conditions where high-quality, annotated images are rare or for pathologies that present with a high degree of variability.

The application of synthetic data in medical image segmentation promises not only to enhance the accuracy and reliability of preoperative planning but also to facilitate the broader adoption of deep learning technologies in clinical settings. As synthetic data generation techniques become more sophisticated, their integration into neurosurgical research and practice is expected to accelerate, offering new opportunities for innovation and improvement in patient care.

### 3.2 Registration and deformation models for surgical planning

Traditionally, the accuracy of surgical planning has been hampered by the limitations of rigid alignment techniques, which struggle to accommodate the anatomical changes that occur during surgery. The challenge of capturing and compensating for these deformations has been a significant hurdle, primarily due to the scarcity of labeled deformation sequences that accurately represent the wide range of tissue dynamics encountered in practice.

Recent advancements, as demonstrated by Han et al. (2022) and Morin et al., showcase the potential of synthetic data to revolutionize this aspect of neurosurgical planning. Han et al. have achieved remarkable registration performance through a joint synthesis and translation network, pre-trained on simulated MRI deformations. This approach not only enhances the accuracy of image registration but also presents a novel methodology for acquiring representative ground truths, bypassing the need for extensive real-world data collection.

Similarly, Morin et al.’s work with polyaffine biomechanical modeling, underpinned by ultrasound scans synthesized from various vantage points, exemplifies the utility of synthetic digital twins in modeling surgical interventions. By simulating deformation trajectories, their approach provides a robust framework for planning and executing neurosurgical procedures, significantly reducing the dependency on immediate intraoperative imaging and the logistical challenges it entails.

These pioneering studies underline the versatility and efficacy of synthetic data and advanced modeling techniques in addressing the complexities of neurosurgical planning. By enabling more accurate predictions of tissue deformation and facilitating the alignment of surgical tools with the operative field, synthetic data is setting new standards for precision in neurosurgery.

The implications of these advancements extend beyond the immediate improvements in surgical accuracy. They herald a new era in neurosurgical preparation, where the reliance on conventional imaging and manual adjustments is replaced by automated, data-driven systems capable of adapting to the dynamic surgical environment. This shift not only promises to enhance the safety and effectiveness of neurosurgical interventions but also opens the door to more innovative approaches to surgery, where the limits of what can be planned and executed are continually expanding.

### 3.3 Retrospective analyses and epidemiological insights

The quest for advancing neurosurgery through data-driven insights often encounters the twin barriers of data accessibility and privacy, especially when attempting to amalgamate heterogeneous records across institutional boundaries for large-scale evidence. In this context, the advent of synthetic data emerges as a beacon of innovation, offering a method to circumnavigate these barriers while preserving the integrity and utility of the aggregated evidence.

Synthetic data’s utility in bridging the gap between the need for comprehensive data and the imperative of confidentiality is exemplified in recent explorations. The work of Khalaf et al. (2023) and Greenberg et al. (2022) serves as a testament to the feasibility and reliability of synthetic datasets in mimicking real-world clinical outcomes. Khalaf et al. (2023) venture into the realm of brain tumor prognostic studies, employing synthetic data to accurately replicate key survival predictors. Their findings not only validate the effectiveness of synthetic datasets in echoing real-world statistical properties but also underscore the potential for these tools to evolve alongside scientific advancements, thereby enhancing the granularity and accuracy of disease progression models.

Parallelly, Greenberg et al. (2022) contribute to the discourse by comparing real and synthetic datasets within the domain of spine surgery. Their investigation reveals a striking similarity in descriptive characteristics and predictive model estimates concerning surgical complications, further reinforcing the notion that synthetic data can serve as a viable stand-in for actual patient data. This alignment between synthetic and real datasets illustrates the robustness of synthetic data in facilitating multicenter studies, which are often hampered by logistical and governance constraints.

Together, these studies illuminate the path forward for neurosurgical research, advocating for a paradigm shift towards the integration of synthetic data. By successfully emulating properties of real-world evidence pools, synthetic data not only promises to enhance research access and capacity but also opens the door to a new era of epidemiological insights and retrospective analyses unbounded by the traditional constraints of data sharing norms. The implications of this shift are profound, offering a scaffold upon which future neurosurgical advancements can be built, free from the encumbrances of privacy concerns and data scarcity.

### 3.4 Accessing multi-center data for research

The landscape of neurosurgery research is often hindered by the difficulty of pooling data from various institutional repositories into unified datasets that are both statistically robust and diverse. This challenge is compounded by the stringent requirements of ethical data governance, which mandate the protection of patient confidentiality and the responsible use of sensitive information. Within this complex framework, the advent of deidentified synthetic data presents a promising avenue for surmounting these obstacles, offering a method that respects privacy concerns while facilitating comprehensive research endeavors.

Recent explorations into this domain, notably by Greenberg et al. (2022) and Khalaf et al. (2023), underscore the significant potential of synthetic data to mimic the intricate details of real patient datasets across multiple healthcare settings. Greenberg et al. (2022) demonstrate that sophisticated algorithms can generate synthetic datasets that closely resemble the descriptive properties and predictive relationships found in pooled electronic health records, thereby suggesting a path forward for multi-center studies that have traditionally been bogged down by privacy and data ownership issues. Their advocacy for the creation of national synthetic data repositories speaks to the transformative impact such a resource could have on facilitating research across institutional boundaries, eliminating traditional barriers to data sharing.

Concurrently, Khalaf et al. (2023) address a critical aspect of synthetic data generation—the representation of rare subpopulations. Their work acknowledges existing limitations in this area but also highlights the dynamic nature of synthetic data methodologies, which can evolve to incorporate new findings and insights from a broad spectrum of literature. This adaptability ensures that synthetic data can remain a relevant and powerful tool for neurosurgical research, particularly in studies that require nuanced understandings of less common conditions or patient demographics.

The contributions of these studies lay the groundwork for a new paradigm in neurosurgical research, one that leverages synthetic data to bridge the divide between the need for comprehensive, heterogeneous datasets and the imperatives of ethical data use. By moving towards standardized tools and protocols that emphasize the role of expert oversight in the generation and application of synthetic data, the neurosurgery research community can make significant strides in advancing our understanding of neurological conditions and treatments. This shift not only promises to expand access to and diversity of research data but also reinforces the commitment to conducting research in a manner that is both ethically sound and scientifically robust.

### 3.5 Privacy-preserving analytics and predictive modeling

The integration of analytics into neurosurgery, leveraging diverse patient data, holds immense potential for refining clinical decision-making processes. However, the ethical imperative to protect patient privacy requires that any data used in such analyses be thoroughly de-identified, posing a significant challenge to maintaining the utility of these datasets for predictive modeling. In this landscape, synthetic data emerges as a critical solution, adept at preserving the intricate predictive relationships characteristic of original datasets while ensuring absolute anonymity.

Illustrating this potential, Greenberg et al. (2022) delve into the application of synthetic data for risk stratification in neurosurgery, demonstrating that models trained on synthetic datasets exhibit minimal deviation in performance compared to those trained on real-world data, particularly in predicting surgical complications. This finding not only validates the efficacy of synthetic data in maintaining the predictive quality of original datasets but also highlights its utility in circumventing privacy concerns that often restrict data usability.

Complementing this, Schonfeld et al. (2023) explore the generation of synthetic datasets designed to enhance the diversity of training cases for predictive models without breaching privacy protocols. By enabling the inclusion of a broader array of clinical scenarios, these synthetic datasets facilitate a more comprehensive understanding of patient outcomes, thereby broadening the scope of research questions that can be addressed within the constraints of privacy regulations.

Furthermore, Khalaf et al. (2023) extend the application of synthetic data to the realm of prognostic modeling, successfully replicating time-to-event models that offer comparable survival risk classifications to those derived from actual patient histories, without compromising sensitive information. This achievement not only showcases the versatility of synthetic data in supporting a wide range of analytical needs but also reinforces its role in fostering responsible research practices that respect patient confidentiality.

Together, these studies underscore a paradigm shift towards the adoption of synthetic data in neurosurgery research, advocating for a model of data analysis that adheres to the highest standards of privacy and ethical responsibility. By enabling the generation of rich, diverse, and privacy-compliant datasets, synthetic data offers a pathway to advance neurosurgical analytics and predictive modeling, ensuring that research can progress without compromising the trust and safety of patient populations. This approach not only aligns with the ethical obligations of the medical research community but also sets a precedent for the development of Shareable, Aggregatable, Reusable, and Private (SHARP) data ecosystems tailored to the nuanced needs of neurosurgery and beyond.

### 3.6 Synthetic CT scans for surgical planning and neuronavigation

In the domain of neurosurgery, the precision of surgical interventions is paramount, heavily relying on the detailed anatomical insights provided by preoperative imaging. While MRI is unparalleled in soft tissue visualization, the integration of CT data is indispensable for defining surgical pathways and entry points with the utmost accuracy. However, the requisite repeated CT scans expose patients to additional radiation, presenting a dilemma between achieving surgical precision and minimizing patient risk.

Addressing this challenge, Staartjes et al. (2021) present a groundbreaking approach through the application of deep learning techniques to synthesize CT images from routine MRI scans. This method not only circumvents the need for multiple CT exposures but also retains the high fidelity necessary for accurate surgical planning and neuronavigation. The synthetic CT maps generated through this process are demonstrated to preserve the essential target registration accuracy, thus supporting the critical requirements of neuronavigation systems without the associated radiation risk.

The significance of this advancement extends beyond the technical achievement; it represents a pivotal shift towards more ethical surgical practices. By leveraging synthetic CT scans, neurosurgeons can refine surgical trajectories and plan interventions with reduced patient exposure to radiation, particularly benefiting procedures like spine stabilization where precision is crucial but the cumulative radiation dose from repeated imaging raises ethical concerns.

Staartjes et al. (2021) underscore the potential of synthetic data to harmonize the objectives of surgical precision and patient safety. This innovative approach not only streamlines the planning phase of neurosurgical procedures but also sets a new standard for responsible medical practice, emphasizing the importance of minimizing patient risk without compromising the quality of care.

### 3.7 Augmenting visualization with expected appearances

The precision of image-guided neurosurgery is paramount, requiring accurate alignment between pre-operative plans and real-time anatomical landmarks. The dynamic nature of surgical environments, characterized by tissue shifts and perspective changes, often complicates this alignment, introducing potential risks to patient safety. Addressing these challenges, synthetic data emerges as a revolutionary tool, offering a method to predict and visualize anatomical changes during surgery, thereby enhancing the accuracy of neuronavigation.

Haouchine et al. (2023) spearhead this innovation with their development of a framework capable of synthesizing photo-realistic images that predict the appearance of surgical scenes. Grounded in physics-based modeling and leveraging baseline patient scans, their approach generates anticipated visual cues that align with intraoperative reality. This technique allows for the adjustment of neuronavigation systems in real-time, accommodating tissue deformations and shifts that occur during the surgical process. By providing surgeons with a more accurate representation of the operative field, these synthetic visual aids significantly bolster the reliability of intraoperative navigation, ensuring that surgical interventions are both safe and effective.

The implications of this technology extend beyond the immediate enhancement of surgical precision. By improving the fidelity of intraoperative visualization, synthetic data-based frameworks like that of Haouchine et al. (2023) contribute to a broader sense of assurance among surgical teams. This confidence is critical during complex or high-risk procedures, where the margin for error is minimal, and the consequences of inaccuracies can be significant. Moreover, the ability to accurately predict and visualize surgical outcomes facilitates more informed decision-making in the operating room, potentially reducing the incidence of complications and improving patient outcomes.

### 3.8 Biomechanical modeling and ultrasound integration

The quest for precision in neurosurgery often grapples with the complexity of anatomical deformations occurring during procedures. Traditional methods, while effective, introduce interruptions in the surgical workflow due to the need for intermittent imaging. This challenge has spurred the development of innovative solutions that combine real-time imaging modalities, such as intraoperative ultrasound, with advanced biomechanical modeling techniques. These integrations are particularly promising for their ability to offer continuous guidance without the procedural pauses typically necessitated by conventional imaging.

A pioneering study by Morin et al. exemplifies this innovative approach, showcasing how ultrasound data can inform finite element models to simulate brain tissue and vascular movements accurately. This synergy allows for the real-time prediction of parenchymal shifts and deformation, facilitating a dynamic adaptation of surgical plans in response to observed changes. By leveraging synthetic data to refine and stabilize simulation parameters across a multitude of procedural scenarios, the model achieves a high degree of accuracy in forecasting anatomical changes, thus ensuring surgical precision and safety.

This methodology represents a significant advancement in neurosurgical practices, offering a solution to one of the most persistent challenges in the field: maintaining the fidelity of surgical navigation in the face of inevitable anatomical shifts. The work of Morin et al. underscores the potential of integrating biomechanical modeling with ultrasound, providing a real-time, adaptive framework that can significantly enhance the surgeon’s ability to make informed decisions based on the current state of the anatomy.

Moreover, the application of synthetic data in this context emerges as a crucial enabler, allowing for the refinement of models to a degree previously unattainable. It facilitates a deeper understanding of tissue dynamics and deformation patterns, thereby improving the model’s predictive capabilities. However, the call for broader validation studies highlights the need for further exploration into the generalizability of these models across varied surgical scenarios and anatomical conditions. Addressing these challenges will require a concerted effort to assess the robustness of these integrative approaches against the backdrop of tissue heterogeneity and the complex rheological properties of brain tissue.

### 3.9 Mixed reality surgical rehearsal

The advent of patient-specific rehearsal practices has markedly improved the landscape of preoperative planning and training in neurosurgery. Such rehearsals are crucial for refining surgical techniques and enhancing patient safety, yet the reliance on traditional methods like cadaveric simulations has been fraught with challenges, including high costs, limited availability, and a lack of repeatability and realism. In response to these challenges, recent innovations have leveraged mixed reality technologies and synthetic data to create immersive, high-fidelity virtual environments for surgical training.

A notable advancement in this domain is the development of mixed reality models by Coelho et al., which exemplify the potential of combining holographic visualizations with 3D-printed components. These models are tailored to patient-specific anatomies, offering a realistic platform for device manipulation and scenario rehearsal. By integrating digital visualizations with physical components that mimic tissue haptics, this approach facilitates a comprehensive training experience that closely mirrors the complexities of actual surgical procedures. The authors demonstrate that such mixed reality simulations can significantly enhance trainee performance, providing a more effective and engaging learning experience compared to traditional training methods.

The success of these mixed reality simulations lies in their ability to synthesize the benefits of digital efficiency with the tactile feedback of real-world interactions. This hybrid model ensures that trainees can practice surgical techniques in a controlled, repeatable environment without the logistical burdens and ethical concerns associated with cadaveric specimens. Moreover, the use of synthetic data within these simulations allows for a wide range of scenarios to be replicated, from routine procedures to rare and complex cases, thus broadening the scope of surgical education.

Despite the promising outcomes associated with mixed reality surgical rehearsal, the need for ongoing refinement, particularly in enhancing dynamic feedback mechanisms, remains. Addressing these areas of improvement will further optimize the realism and educational value of these simulations, ensuring that they can accurately replicate the nuances of surgical interventions.

### 3.10 Patient-specific rehearsal with 3D printing

Neurosurgical interventions demand a precise understanding of patient-specific anatomies, often too complex to be fully grasped through two-dimensional imaging alone. Addressing this challenge, recent technological advancements have harnessed the power of synthetic data to create detailed 3D-printed models that replicate the unique anatomical features of individual patients. These models serve as a bridge between theoretical knowledge and practical application, offering a tangible means for surgeons to rehearse and refine their techniques on patient-specific anatomies before actual surgery.

A pioneering example of this application is the work of Licci et al., who have developed reusable phantom heads designed to simulate various brain tumor scenarios. These models incorporate interchangeable parts that mimic the physical properties of tumors, including texture variations that are customizable to match the specific imaging characteristics of patient cases. By utilizing a comprehensive dataset to inform the variability in tumor presentation, the models achieve a balance between realism and variability, thereby enhancing the educational value of surgical rehearsals.

The introduction of these 3D-printed models into surgical training programs has been met with positive feedback from trainees, who particularly value the opportunity to practice navigating complex cranial structures through transparent access corridors designed into the phantoms. This aspect of the training aids in building confidence and competence in handling a range of surgical scenarios, from routine to highly complex interventions.

The significance of this innovation extends beyond the immediate benefits of enhanced surgical training. By providing a means to visualize and interact with patient-specific anatomies in a controlled setting, 3D-printed models augmented with synthetic data offer a unique opportunity to anticipate and strategize around potential challenges that may arise during actual surgical procedures. This preparatory tool is invaluable in improving surgical outcomes, reducing operative times, and enhancing patient safety.

### 3.11 Validating training models and metrics

The quest for effective neurosurgical training methodologies increasingly turns towards simulation-based environments, which promise a risk-free setting for developing and honing surgical skills. Central to the utility of these simulations is the establishment of reliable metrics that can accurately measure a trainee’s competence, predicting their ability to translate simulated practice into real-world surgical success. The challenge lies in validating these metrics against actual surgical outcomes, a task complicated by the intricate nature of neurosurgical procedures and the ethical implications of direct experimentation on patients.

Innovative research, such as the work conducted by Ahmed et al., showcases the potential of synthetic simulators to bridge this gap. By employing a synthetic aneurysm model for aneurysm clipping rehearsals, the study evaluates the validity of simulation metrics across different levels of surgical expertise. Through detailed motion analytics, significant differences in performance metrics are identified among novice, intermediate, and expert surgeons, underscoring the simulator’s ability to distinguish between varying levels of skill. This differentiation is crucial for the development of a competency-based training framework that can reliably predict a surgeon’s readiness for real-life procedures.

Moreover, the findings from such studies underscore the importance of continuous innovation in simulation technology, particularly in the materials and methods used to replicate the physical and physiological characteristics of human anatomy. By enhancing the realism and biofidelity of synthetic models, training simulations can provide even more accurate assessments of surgical skill, further closing the gap between simulated practice and actual operative performance.

The implications of this research extend beyond the validation of training metrics; they also highlight the potential of synthetic simulators as a platform for developing advanced assessment protocols. These protocols could revolutionize how surgical competency is measured and certified, shifting towards a more objective and quantifiable standard that directly correlates with patient outcomes.

### 3.12 Other applications

The assimilation of synthetic data and artificial intelligence (AI) into neurosurgery heralds a pivotal shift, poised to redefine conventional methodologies spanning from preoperative strategizing to post-operative management. This systematic review amalgamates insights from a curated collection of studies, shedding light on the expanding influence of these innovations in neurosurgical progress, and accentuating their multifaceted impact on the discipline.

In the domain of surgical planning and execution, the pioneering application of synthetic data in assessing optical flow methods, as delineated by Markus Philipp et al. (2021), signifies a substantial advancement. Their endeavors, focused on synthesizing neurosurgical microscope imagery, highlight the indispensable role of synthetic data in algorithmic evaluation and benchmarking. This methodology not only replicates the intricacies of actual neurosurgical settings but also facilitates a detailed assessment of optical flow algorithms, with PWC-Net demonstrating superiority over the Farneback method. Such progress indicates a route towards refining computational instruments, potentially elevating intraoperative precision and impacting patient safety and surgical results positively.

Concurrently, the synergy of AI and connectomics in neurosurgery, explored by Antonio Di Ieva et al. (2021), unveils the potential of deep learning models in transforming brain tumor surgeries. Achieving remarkable accuracy in tumor segmentation and genetic subtype discernment, this integration furthers personalized treatment approaches, thereby significantly enhancing surgical planning accuracy and aligning with the overarching aim of customizing interventions to match individual patient needs, a cornerstone of the precision medicine approach in neurosurgery.

Moreover, the comparative study of synthetic versus autologous dura mater in pediatric neurosurgery by Emir Kaan İzci et al. (2023) offers a discerning view on material selection and its repercussions on postoperative outcomes. This analysis underscores the critical need for judicious material choice, advocating for synthetic data’s utility in guiding clinical decision-making and potentially steering innovations in surgical materials to bolster patient results.

Additionally, the foray into synthetic learning for spine surgery by Ethan Schonfeld and Anand Veeravagu (2023) tackles the challenge posed by the scarcity of extensive datasets, attributed to privacy and data sharing restrictions. The creation of SpineGAN for synthetic radiograph generation exemplifies how synthetic data can surmount traditional hindrances, promoting enhanced model training while safeguarding patient confidentiality, indicative of a wider trend towards ethical medical research enhancement through synthetic data and AI.

Lastly, advancements in intraoperative registration techniques by Nazim Haouchine et al. (2023), through the use of AI in synthesizing preoperative expected appearances, mark a significant stride in surgical navigation. This methodological innovation, by bolstering the alignment accuracy between preoperative MRI and intraoperative visuals, not only refines neurosurgical navigation fidelity but also pioneers the integration of AI solutions into real-time surgical procedures, heralding improved patient care standards and safety.

## 4 Discussion

### 4.1 Summary of evidence

The systematic review explores the critical role of data in the evolution of neurosurgery, highlighting the innovative applications of synthetic data throughout the surgical process, from preoperative planning to postoperative care. The shift towards personalized medicine is exemplified through the use of patient-specific anatomical models and predictive analytics, enabling surgeons to devise tailored surgical interventions. Intraoperative navigation systems, leveraging multimodal imaging and biomechanical models, provide real-time guidance, improving surgical accuracy and safety by accounting for anatomical shifts during operations. In the postoperative phase, data analytics play a pivotal role in shaping individual recovery plans, signifying a move towards a more data-centric approach in patient management.

The review acknowledges the challenges posed by the limited availability and variable quality of clinical data, presenting synthetic data generation and simulation as viable solutions. Synthetic data stands out for its capacity to mimic real-world data’s statistical properties, offering a flexible resource that supports research, training, and the enhancement of surgical procedures, unfettered by the limitations of conventional data sources.

Prominent research, including works by Lindner et al. (2019), Pantovic et al. (2022), Han et al. (2022), and Morin et al., demonstrates the effectiveness of synthetic data in addressing specific neurosurgical challenges—from refining CT scan analysis to improving surgical planning with sophisticated modeling techniques. The utility of synthetic data in generating real-world evidence for retrospective studies and epidemiological research, as shown by Khalaf et al. (2023) and Greenberg et al. (2022), highlights its capacity to reconcile the demand for extensive data with privacy considerations.

Furthermore, the application of synthetic data in surgical assistance and planning, notably through the generation of synthetic CT scans for neuronavigation and the enhancement of surgical visualizations, marks a significant advancement. Innovations such as mixed reality surgical rehearsals and 3D-printed models for patient-specific practice underscore synthetic data’s contribution to elevating surgical training and preparation.

Yet, translating these theoretical models into practical applications presents hurdles, such as achieving a balance between the realism and diversity in synthetic data and integrating these technologies into clinical practice. Overcoming these obstacles will require continuous improvement, cross-disciplinary collaboration, and a nuanced approach to ensure the successful integration of synthetic data applications that are both technologically sophisticated and clinically applicable.

### 4.2 Future directions for research

The burgeoning field of synthetic data science offers promising avenues for revolutionizing neurosurgical practices, touching every aspect of the data value chain—from patient-specific planning to global health trend analysis. While current endeavors have shown significant promise through experimental studies within specialized centers, the broader application of these technologies necessitates a concerted push towards practical, applied uses that directly benefit patient outcomes and public health at large.

A pivotal step in this journey is the development of decentralized, yet responsibly managed, data repositories. As suggested by Pandit et al., such frameworks could vastly improve the scope and quality of analytics at a national level, uncovering population health trends that inform strategic policymaking and resource allocation. However, realizing this potential hinges on overcoming the cultural inertia that currently pervades medical data practices. This calls for a paradigm shift in how medical data is accredited, managed, and utilized, emphasizing skills development and the adoption of new technologies.

Central to achieving this vision is the establishment of data cooperatives, interconnected through standardized APIs, fostering a culture of continuous improvement and collaboration among a wide range of stakeholders. By credentialing these cooperatives, the neurosurgical community can ensure a steady flow of high-quality, standardized data, fueling innovations that directly address patient needs and healthcare challenges.

Moreover, adopting a more inclusive approach to development is crucial for ensuring that the benefits of synthetic data reach all corners of the globe. By amplifying the voices of marginalized communities and incorporating diverse perspectives into the development process, the neurosurgical field can create more representative and effective simulation models. This inclusivity extends to the pooling of learning resources and the propagation of analytics tools that are both locally relevant and globally applicable, ensuring that advancements in synthetic data science translate into tangible improvements in patient care.

### 4.3 Integration into clinical workflows

The potential of synthetic data to revolutionize neurosurgical practices through high-fidelity simulations and patient-specific planning is increasingly evident. Experimental validations have showcased the capability of these data paradigms to overcome the constraints posed by traditional data sources, offering a glimpse into a future of more informed and accurate surgical interventions. However, the leap from theoretical feasibility to routine clinical application is fraught with hurdles, primarily revolving around the seamless integration of synthetic data technologies into existing neurosurgical workflows.

Illustrative of these challenges are the observations by Haouchine et al., who acknowledge that while their registration algorithms demonstrate promising utility in offline settings, incorporating these technologies into live, image-guided surgical navigation systems—complete with real-time data processing—remains unachieved. This discrepancy between experimental capability and practical usability underscores the necessity of adapting synthetic data tools to fit within the complex ecosystem of surgical hardware and software, ensuring that they enhance rather than disrupt the surgical process.

Similarly, the vision articulated by Greenberg et al. for creating national-scale, aggregated synthetic datasets capable of supporting multicenter trials hinges on the establishment of unified data standards and interoperability protocols. Such an undertaking necessitates not just technological innovation but also a concerted effort to harmonize data structures across diverse healthcare and research platforms, ensuring that synthetic data can be seamlessly integrated and utilized across various settings.

These examples highlight a critical gap in the current trajectory of synthetic data integration into neurosurgery: the need for a multidisciplinary approach that bridges computational science, engineering, and clinical practice. Effective translation of synthetic data technologies into clinical settings requires the active involvement of cross-functional teams dedicated to co-creating solutions that are not only technologically advanced but also attuned to the practical realities of surgical care. This collaborative model, rooted in the principles of Responsible Research and Innovation, is essential for developing tools that are both usable in real-world settings and capable of contributing meaningally to patient outcomes.

Addressing the integration challenge also calls for a reevaluation of existing clinical workflows, identifying opportunities to embed synthetic data tools in a manner that complements and enhances surgical planning and execution. By focusing on creating intuitive, user-friendly interfaces and ensuring compatibility with existing surgical equipment, synthetic data applications can become a natural extension of the neurosurgeon’s toolkit.

### 4.4 Limitations and Challenges

The potential of synthetic data to revolutionize neurosurgical practice is immense, offering unprecedented opportunities for training, rehearsal, and research. Yet, as we delve deeper into its application, the intricacies of achieving an optimal balance between the realism of specific domain features and the diversity of surgical scenarios come to the forefront. A critical examination of existing models reveals areas where current synthetic data generation techniques fall short, particularly in capturing the full complexity of neurosurgical procedures.

For instance, the work by Staartjes et al. highlights a limitation within MRI-derived synthetic CT models, which accurately replicate anatomical shapes but fail to account for variations in bone density—a critical factor for surgical assessments involving drilling. This omission points to a gap in the models’ ability to fully prepare surgeons for the tactile experiences encountered during actual procedures.

Similarly, Licci et al. have identified areas for improvement in the realism of tumor simulations within phantom heads, specifically regarding texture cues and the emulation of bleeding. These aspects are vital for accurately representing the heterogeneity of pathological conditions encountered in neurosurgery. Enhancing these features would not only improve the educational value of such models but also their applicability in planning and rehearsing complex surgical interventions.

These examples underscore the nuanced challenge of designing synthetic data models that accurately reflect the wide range of variables inherent in neurosurgical procedures. The risk of overfitting models to specific scenarios or features—thereby limiting their utility in unfamiliar or varied surgical contexts—highlights the need for a comprehensive approach to synthetic data generation. This approach must incorporate a purpose-driven analysis of requirements, ensuring that models achieve a harmonious balance between case-specific detail and the broader distribution of features encountered across neurosurgical practices.

Addressing these challenges requires a concerted effort to refine synthetic data generation processes, integrating advanced generative techniques that can capture the complexity and variability of the neurosurgical field. By achieving this balance, synthetic data can fulfill its promise as a transformative tool in neurosurgery, enhancing training, planning, and execution of procedures with unprecedented precision and reliability.

## 5 Conclusions

The evolution of synthetic data generation in neurosurgery heralds a new era of data-driven advancements, addressing longstanding limitations that have constrained the field’s progress. Recent developments showcase an array of synthetic data applications, each designed to enhance different facets of neurosurgical care. From creating detailed simulations of tumor imagery for preoperative planning to facilitating mixed reality environments for surgical training, these innovations are setting new standards for precision and patient-specific care.

Central to the appeal of synthetic data is its ability to generate realistic, detailed proxies for various neurosurgical needs, significantly reducing reliance on real-world datasets, which are often limited in availability and scope. This capability is crucial for advancing neurosurgical practices, offering a rich resource for training, research, and procedural planning. For example, the initiatives by Greenberg et al. to establish national-scale synthetic hospital repositories exemplify the potential to democratize access to comprehensive data sets, enabling more inclusive research and development activities across the neurosurgery community.

These synthetic data solutions not only promise to enhance the accuracy and effectiveness of current neurosurgical protocols but also aim to streamline the integration of artificial intelligence into clinical workflows. By providing a consistent, readily available source of data for AI training, synthetic datasets facilitate the development of robust, deep learning algorithms capable of supporting a wide range of neurosurgical applications, from diagnostic assistance to personalized treatment planning.

However, the journey towards fully realizing the potential of synthetic data in neurosurgery is not without its challenges. Issues such as ensuring representation of rare subpopulations, harmonizing protocols across multiple centers, and seamlessly integrating synthetic data tools into clinical workflows necessitate ongoing refinement and collaboration. Addressing these concerns is essential for fostering the development of synthetic data applications that are not only technologically advanced but also clinically relevant and widely accessible.

## Supporting information

Systematic Review Matrix

PRISMA Search Results

## Data Availability

All data produced in the present study are available upon reasonable request to the authors

