## Supplementary material for "The Emerging Applications of Synthetic Data in Neurosurgery Research and Practice: A Systematic Review": Systematic Review Matrix

| Author/ Date | Theoretical/ Conceptual Framework | Research Question(s)/ Hypotheses | Methodology | Analysis & Results | Conclusions | Implications for Future research | Implications For practice |
|---|---|---|---|---|---|---|---|
| Jacob K. Greenberg et al., 2022 | Validating the use of synthetic data derivatives for spine surgery research using electronic health records. | Can synthetic data accurately replicate real data for describing outcomes and building predictive models in spine surgery populations? | Compared descriptive statistics and predictive modeling using real EHR data vs. synthetic data for two spine surgery cohorts - anterior cervical fusion (n=9,072) and posterior lumbar fusion (n=12, 111). Predictive outcomes were 30-day hospital readmissions and complications. | Descriptive characteristics nearly identical between real and synthetic datasets. Predictive discrimination was similar using regression and machine learning in both cohorts - C-statistics ranged 0.66-0.71 for readmissions, 0.74-0.86 for complications. Some differences in influential variables. | Synthetic data derivatives effectively simulated most descriptive and predictive properties of real spine surgery data. | Expand validation to additional spine surgery populations; evaluate impact of enhancing EHR data elements. | Synthetic data can enable multicenter spine research by overcoming privacy barriers and data ownership concerns. |
| Roy Khalaf et al., 2023 | Validating the use of synthetic data to reproduce findings from published neuro-oncology studies. | Can synthetic data reliably reproduce results from published clinical research studies in neuro-oncology? | Reproduced two published prognostic studies in neuro-oncology using synthetic data derived from an Israeli hospital EHR. Compared survival outcomes and prognostic factors between real and synthetic datasets. | Synthetic data findings showed significant consistency with real data results for prognostic factors and survival outcomes in both studies. Minor variability between synthetic datasets from same query. | Synthetic data can reliably reproduce key statistical characteristics and findings from real-world clinical data in the neuro-oncology domain. | Incorporate additional data types into synthetic data generation like genotypes, imaging, disease progression. Collaborate with multiple institutions. | Synthetic data enables clinical research while protecting patient privacy. Could be used for hypothesis testing, epidemiological predictions, and education in neuro-oncology. |
| Lydia Lindner et al., 2019 | Using synthetic training data for deep learning-based segmentation of glioblastoma (GBM) in MRI images. | Can a convolutional neural network be successfully trained on synthetic data to segment GBM tumors? | Generated synthetic GBM MRI images and ground truth labels. Trained a U-Net only on synthetic images. Evaluated on real patient MRIs using segmentation metrics like Dice score. | Model achieved 86.2% Dice score on test set. Qualitative assessment showed accurate segmentation for tumors with clear margins. Difficulty with diffuse tumors. | Deep learning GBM segmentation is feasible using solely synthetic training data. | Enhance realism of synthetic tumors. Apply approach to other cancer types. | Synthetic data can enable training models despite lack of real annotated medical images. Could assist diagnosis and treatment planning. |
| Anja Pantovic et al., 2022 | Using synthetic data to train deep learning model for segmentation of SEEG electrodes in post-operative CT scans. | Can synthetic data improve segmentation accuracy compared to only real or augmented data? | Generated synthetic CT volumes with simulated SEEG electrodes and metal artifacts. Trained UNet on real data, augmented data, synthetic data, and combinations. Compared segmentation performance. | Adding synthetic data significantly improved detection of contacts compared to real or augmented data alone, with higher Dice score, IoU, TPR, and PPV. | Incorporating synthetic data during training enables more robust SEEG electrode segmentation in the presence of metal artifacts. | Address limitations like merged contacts on perpendicular electrodes. | Synthetic data can reduce human error and save time in localizing SEEG electrodes for epilepsy surgery. |
| Victor E. Staartjes et al., 2021 | Utilization of MRI-based synthetic CT (sCT) for spinal neuronavigation to mitigate the need for CT scans in spinal surgery planning. Deep learning approaches for generating sCT from MRI images. | Evaluate the feasibility and effectiveness of MRI-based sCT for lumbar spine surgical planning and its potential to replace conventional CT scans. | Synthetic CT reconstructions using "BoneMRI" software, relying on a deep learning–based image synthesis method trained on paired MRI-CT data. | sCT allowed for qualitatively adequate CT images of the lumbar spine, showing normal and pathological structures reliably. | The study demonstrates the potential of MRI-based sCT in reducing workflow complexity, radiation exposure, and costs, while maintaining adequate quality for surgical planning and neuronavigation. | Further validation of the method in patients with various spinal conditions and implants. Development based on larger patient cohorts to improve image fidelity. | This novel method could revolutionize navigated lumbar spine surgery by eliminating the need for radiation-exposure through CT scans, simplifying surgical planning and potentially improving patient outcomes. |
| Perrine Paul, Xavier Morandi, Pierre Jannin, 2009 | The study focuses on quantifying intraoperative brain deformations in image-guided neurosurgery (IGNS) to improve surgical outcomes. | Investigates the feasibility and accuracy of a new surface registration method for quantifying brain deformations using textured cortical surface meshes and video flow. | Utilizes a nonrigid registration method combining geometric information, texture matching, and sparse landmarks matching for tracking cortical surface deformations. | Demonstrated feasibility in clinical settings with precision around 2 mm, comparable to usual rigid registration errors in neuronavigation systems before deformations. | Highlights the method's potential for real-time, accurate tracking of brain deformations, enhancing the effectiveness of IGNS. | Suggests further validation with more clinical cases and the development of dynamic models adapting to surgical steps. | Offers a promising approach for real-time, accurate brain deformation tracking, potentially improving surgical precision and outcomes in neurosurgery. |
| Maria Licci, Florian M. Thieringer, Raphael Guzman, Jehuda Soleman, 2020 | The need for realistic, cost-effective, and reusable surgical simulators for training in neuroendoscopic ventricular lesion removal due to limitations in current surgical education methods. | To develop and validate a patient-specific, low-cost, 3D-printed simulator for neuroendoscopic ventricular lesion removal training and assess its realism, mechanical properties, procedural content, and handling. | Development of a synthetic simulator based on patient-specific CT data, including realistic skull models and a tumor model from polyvinyl alcohol, assessed by neurosurgical trainees through a feedback survey. | High appreciation for the model as a valid training tool, with realistic anatomical and mechanical properties, indicating potential improvements like different tumor textures and realistic surgical approaches. | Successful development of a patient-specific, reusable 3D-printed simulator for neuroendoscopic ultrasonic aspirator tumor removal training, recognized as a useful educational tool. | Further development of different tumor model textures and user ability to create realistic surgical skull approaches; expansion to include a broader spectrum of neuroendoscopic procedures. | Enhances neurosurgical training by providing a safe, reproducible environment for practice, potentially improving surgical skills and patient outcomes in neuroendoscopy. |
| R Han et al., 2022 | This study develops a deep learning-based deformable registration method to address deformations between preoperative MR and intraoperative CBCT in neurosurgery. | The study investigates the efficacy of a joint image synthesis and registration network (JSR) for addressing brain deformations in neurosurgical guidance. | The JSR method uses a multi-resolution pyramid for image synthesis and registration, initially trained on simulated data and refined on clinical images. | JSR achieved superior registration accuracy with median Dice coefficient (DSC) and median target registration error (TRE) significantly better than other methods, supporting its effectiveness. | JSR resolves deep brain deformations between MR and CBCT images efficiently, offering superior performance over existing state-of-the-art methods. | Further clinical studies are suggested to validate JSR's effectiveness and explore its application in other surgical contexts. | The method enhances the accuracy and reliability of neurosurgical navigation, potentially improving surgical outcomes by better addressing intraoperative brain deformations. |
| Antonio Di Ieva, Carlo Russo, Abdulla Al Suman, Sidong Liu, 2021 | The study explores the intersection of artificial intelligence (AI) and connectomics in neurosurgery, specifically for brain tumor pre- and intra-operative imaging. | The research aims to determine the effectiveness of AI models in segmenting brain tumors on MRI images, predicting genetic subtypes of glioma, and planning resections based on connectomics data. | Utilized deep learning models trained on a dataset of brain tumor images for segmentation, digitalized histopathology slides for genetic subtype prediction, and connectomics data to customize resection strategies. | Achieved high accuracy in tumor segmentation and genetic subtype prediction, demonstrating the utility of AI and connectomics in enhancing surgical planning and outcomes. | The study concludes that AI and connectomics significantly contribute to the precision and safety of brain tumor surgeries, supporting their integration into clinical practice. | Suggests further research on integrating these technologies into real-time surgical navigation systems and exploring their potential in other neurosurgical applications. | Highlights the potential of computational neurosurgery to revolutionize brain tumor management by enabling more accurate diagnostics, personalized surgical planning, and potentially improved patient outcomes. |
| Emir Kaan İzci, Fatih Keskin, Densel Araç, 2023 | This study focuses on comparing synthetic dura mater to autologous dura mater in children aged 0-1 years undergoing surgery for meningocele and myelomeningocele to determine complication risks. | The study aims to evaluate whether the use of synthetic dura mater leads to higher complication rates compared to autologous dura mater in the specified patient population. | A cross-sectional observational study with 44 children, comparing postoperative outcomes between those receiving synthetic versus autologous dura mater. | Synthetic dura mater use was associated with a higher rate of secondary surgery and complications, including infection, necrosis, and neurological deficits, compared to autologous dura mater. | Synthetic dura mater, while necessary in more severe cases, has a higher complication risk and need for secondary surgery than autologous dura. Both primary closure and Limberg flap procedures have similar safety and efficacy in synthetic dura users. | Further research is needed on optimizing dura mater choice and surgical techniques to reduce complications and improve outcomes in neurosurgical pediatric patients. | This study highlights the importance of careful selection between synthetic and autologous dura mater in pediatric neurosurgery, suggesting a preference for autologous dura when possible to reduce complications. |
| Ethan Schonfeld & Anand Veeravagu, 2023 | The study explores synthetic learning in spine surgery for multi-center model training with enhanced patient privacy. It addresses the challenge of limited "big data" in neurosurgery due to patient privacy and data sharing constraints. | Investigates the efficacy of synthetic learning for generating and using synthetic data in spine surgery imaging to train machine learning models without compromising patient privacy. | Utilizes generative adversarial networks (GANs) and a novel SpineGAN for generating synthetic spine radiographs. Machine learning models were then trained on both real and synthetic data to classify spine radiographs as normal or abnormal. | The study achieved successful classification of spine radiographs using synthetic data, with the SpineGAN showing improved performance and diversity in generated images compared to traditional GANs. | Synthetic learning is effective in the spine surgery domain, demonstrating that domain-specific synthetic data generation can enhance model training while preserving patient privacy. | Future research could explore the application of synthetic learning across different medical domains and its integration into clinical practice for broader use. | Highlights the potential of synthetic learning to enable the use of larger, more diverse datasets for training medical imaging models, improving algorithm generalizability while maintaining patient privacy. |
| Nazim Haouchine, Reuben Dorent, Parikshit Juvekar, Erickson Torio, William M. Wells III, Tina Kapur, Alexandra J. Golby, Sarah Frisken, October 3, 2023 | The study introduces a novel approach for intraoperative patient-to-image registration by learning expected appearances using preoperative imaging to synthesize patient-specific expected views. | The research investigates the efficacy of an AI-driven method for synthesizing expected appearances for accurate intraoperative registration in neurosurgery. | Utilized generative adversarial networks (GANs) to synthesize expected appearances from preoperative imaging, and a patient-specific pose regressor network for estimating camera pose by minimizing dissimilarity. | Demonstrated high accuracy in aligning preoperative MRI with intraoperative views, outperforming state-of-the-art methods in both synthetic and clinical data. | Confirms the feasibility and superiority of using expected appearances for intraoperative registration, suggesting a significant advancement in neurosurgical navigation and patient safety. | Suggests exploring non-rigid deformations and adapting to changes in zoom and focal depth during surgery, enhancing texture variability control. | Indicates a shift towards AI-enhanced surgical planning and execution, promising improved accuracy in neuronavigation and broader applications in image-guided surgery. |
| Markus Philipp Neal Bacher, Jonas Nienhaus, Lars Hauptmann, Laura Lang, Anna Alperovich, Marielena Gutt-Will, Andrea Mathis, Stefan Saur, Andreas Raabe, Franziska Mathis-Ullrich, 2021 | The study investigates synthetic data generation for optical flow evaluation in neurosurgery, focusing on the challenge of accurate motion tracking in surgical videos. | It evaluates whether synthetic data can effectively benchmark optical flow algorithms for neurosurgical applications, considering domain-specific visual conditions. | The approach includes generating synthetic neurosurgical microscope data, simulating instrument motion, and evaluating two optical flow algorithms (Farneback and PWC-Net) for accuracy. | The study found that synthetic data effectively supports the evaluation of optical flow algorithms, with PWC-Net outperforming Farneback in accuracy. | Demonstrates the potential of synthetic data in evaluating and improving optical flow algorithms for neurosurgical domain, highlighting the importance of domain-specific challenges in algorithm development. | Future research could explore more complex simulations of neurosurgical environments and the application of synthetic data in training machine learning models for surgery assistance. | Suggests that synthetic data generation can be a valuable tool in developing and benchmarking computational tools for neurosurgery, potentially improving intraoperative guidance and patient outcomes. |
