## Supplementary figures and images for "The Emerging Applications of Synthetic Data in Neurosurgery Research and Practice: A Systematic Review"

### PRISMA Search Results

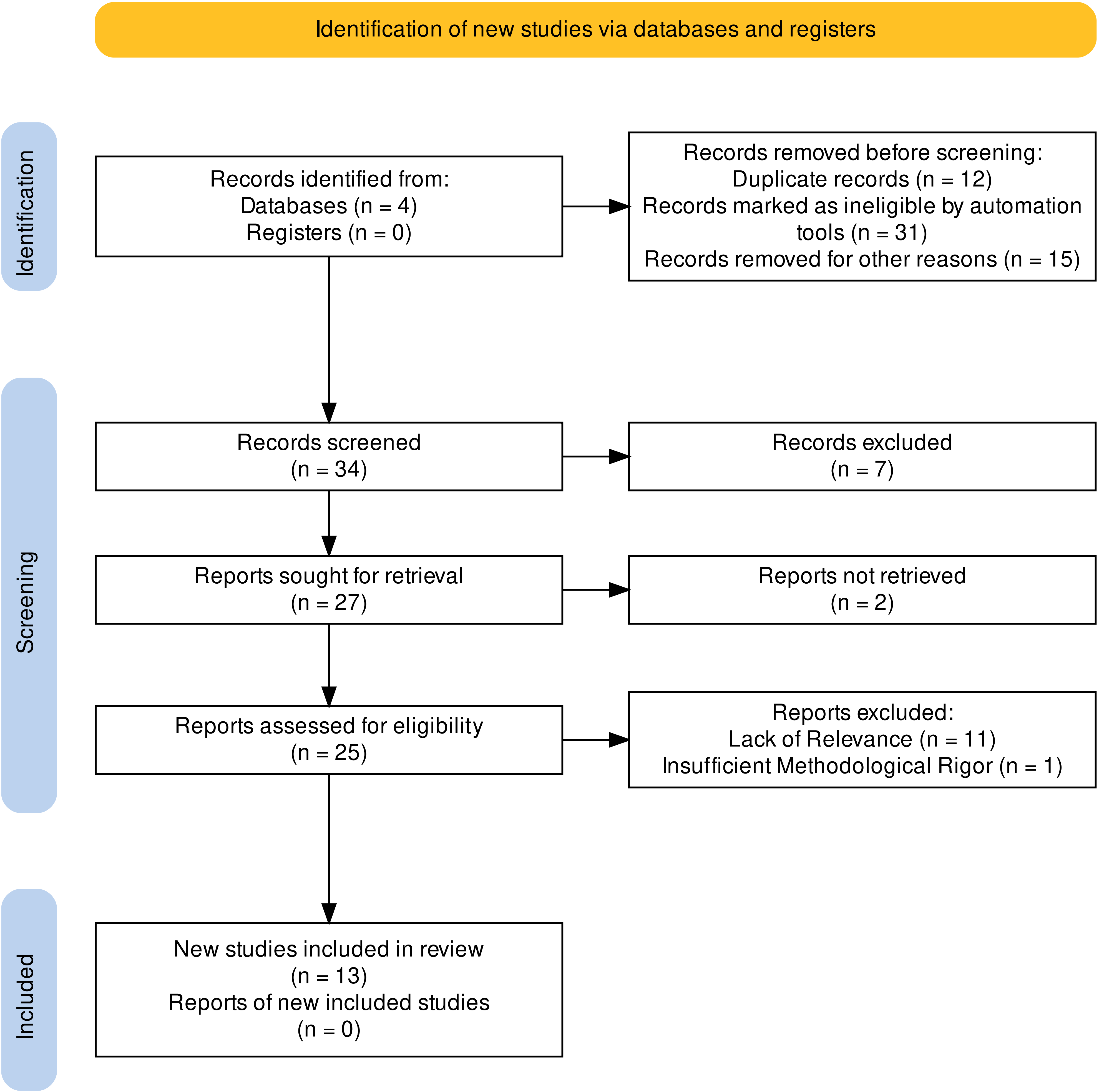
